# Assessing The Relationship Between Demand And Accessibility For Pediatric Ophthalmology Services By State In The United States

**DOI:** 10.1101/2024.08.03.24311451

**Authors:** Benton Chuter, Alexander C. Lieu, Justin Huynh, Jennifer J. Bu, Linda M. Zangwill

**Affiliations:** Hamilton Glaucoma Center, Shiley Eye Institute, Viterbi Family Department of Ophthalmology, University of California San Diego; School of Medicine, University of Illinois Urbana-Champaign, Urbana, IL, United States

**Author notes:** <u>Correspondence to:</u> Benton Chuter, Address: 4240 Porte De Merano, Unit 64, San Diego, CA 92122. <u>Author Statement:</u> **Benton Chuter:** Conceptualization, Methodology, Investigation, Software, Formal Analysis, Writing - Original Draft, Writing - Review & Editing, Visualization **Justin Huynh:** Writing - Review & Editing. **Alexander C. Lieu:** Conceptualization, Methodology, Writing - Review & Editing. **Jennifer J. Bu:** Writing - Review & Editing. **Linda M. Zangwill:** Supervision, Resources Writing- Reviewing and Editing. <u>Disclosures:</u> B. Chuter and AC. Lieu, UCSD MEDGAP 2023. J. Huynh, T35: Short-Term National Research Service Award (NRSA); L.M. Zangwill, National Eye Institute (F), Carl Zeiss Meditec Inc. (F), Heidelberg Engineering GmbH (F), Optovue Inc. (F), Topcon Medical Systems Inc. (F), Zeiss Meditec (P), AISight Health (P), Abbvie (C), Digital Diagnostics (C). LM. Zangwill has received grants from The Glaucoma Foundation, The National Institutes of Health, The National Eye Institute; grants and nonfinancial support from Heidelberg Engineering; nonfinancial support (equipment) from Carl Zeiss Meditec, Optovue, and Topcon. Dr. Zangwill has received consulting fees from AbbVie and Topcon Medical Systems. Dr. Zangwill is co-founder, inventor and board member and equity holder of AISight Health Inc.

## Abstract

**Purpose:** To investigate the relationship between public demand for pediatric ophthalmology services and the accessibility of such services on a statewide basis in the United States, focusing on strabismus care.

**Methods:** Using Google Trends data, search volumes for “strabismus” were analyzed from January 2014 to December 2023. Pediatric ophthalmologist availability was assessed via the American Academy of Ophthalmology directory, normalized for state population size to create a relative demand index. Additional metrics from the United States Census Bureau and Centers for Disease Control and Prevention provided socioeconomic and health data. Correlation coefficients were used to explore relationships between search volumes, specialist availability, vision screening, socioeconomics, and demographic factors.

**Results:** The data revealed variability in public interest in pediatric ophthalmology across states, with Alaska showing the highest relative search volume for strabismus. The data also indicated notable disparities in pediatric ophthalmologist density, with North Dakota, Vermont, and Wyoming having no pediatric ophthalmologists. A significant correlation was identified between pediatric ophthalmologist availability and vision screening (R = 0.486, p < 0.001). Negative correlations were observed between the relative demand index and urbanization (R = −0.388, p = 0.007), as well as the strabismus prevalence (R = −0.455, p = 0.001), indicating that urbanization and specialist distribution influence eyecare demand and access.

**Conclusion:** The findings highlight disparities in accessibility and demand for pediatric ophthalmology services across the US, influenced by urbanization and distribution of healthcare professionals. The study underscores the need for targeted interventions to improve access to care and bridge gaps in pediatric ophthalmology service provision.

## INTRODUCTION

Strabismus, a condition characterized by the misalignment of the eyes, affects approximately 2% to 4% of the population in the United States.^1–6^ Strabismus can lead to amblyopia, loss of depth perception, and social stigma if not treated promptly and effectively.^5,7–12^ Medical care for strabismus may vary based on the severity and type, but surgical intervention to realign the eye muscles remains a common form of treatment.^11,13–17^

Early treatment is crucial for optimal outcomes.^18,19^ Delayed or absent treatment can result in permanent visual impairment, significantly affecting quality of life.^20^ Despite this, delays in treatment are not uncommon due to various factors, including limited access to specialized care, underdiagnosis, and socioeconomic barriers.^19^ Studies suggest that a significant percentage of children with strabismus do not receive timely care, which can exacerbate public health challenges associated with untreated pediatric eye conditions.^21–25^

Lack of availability of pediatric ophthalmologists can contribute to these delays in strabismus care.^26–29^ Availability of pediatric ophthalmology services is critical, especially for conditions like strabismus that require early intervention for optimal outcomes.^30,31^ Pediatric ophthalmologists are relatively underpaid compared to other ophthalmologic specialties, contributing to this shortage.^32,33^ This financial undervaluation can disincentivize pursuit of pediatric ophthalmology as a specialty, further exacerbating the unmet need for specialized eye care for children.^34^

This gap between supply and demand for pediatric ophthalmologist services has been previously explored.^26,28,35^ However, there remains significant uncertainty as to the unmet need for strabismus treatment on a statewide basis, and how that relates to various health and demographic factors. Search volumes for the term “strabismus”, as reported by Google Trends data, may provide additional clarity as to the demand for pediatric ophthalmologists. While Google Trends and other tools have been utilized to understand public interest and awareness in ophthalmology broadly^36^, specific analyses correlating search volumes for strabismus with demographic data and health outcomes as well as pediatric ophthalmologist density on a state-by-state basis has not yet been performed^.37–39^

This work aims to fill this gap by exploring how medical resources align with patterns in population demographics, economic conditions, and specific health metrics across different states as assessed through online search trends data available through Google Trends, pediatric ophthalmologist availability, and demographic and health outcome measures reported by the Census Bureau and the CDC. State-level data regarding ophthalmology and public health metrics is here explored to understand the relationship between ophthalmological services, public health indicators, and demographic factors.

We hypothesize that there are significant disparities in the availability of pediatric ophthalmology services and public demand for strabismus care across different states in the United States, influenced by urbanization, socioeconomic factors, and healthcare professional distribution.

This study could offer valuable insights into the public’s awareness and understanding of strabismus, the demand for healthcare services, and potential barriers to care, contributing to improved public health strategies and resource allocation.

## METHODS

### Data Compilation

To assess demand, Google Trends data was employed. Accessed from trends.google.com, public search query data for the search term “strabismus” from January 1st, 2014, to December 31st, 2023 was acquired as Relative Search Volume (RSV).^40^ The Google Trends platform calculates the RSV by comparing the monthly search frequency for “strabismus” against the total search volume across all states, subsequently averaging these values over the study duration and adjusting them to a 0-100 scale based on relative interest.^41^

To generate data concerning pediatric ophthalmologist availability, the American Academy of Ophthalmology (AAO) online database^42^ was used. The directory’s accuracy was also confirmed by comparing listings with the results given by the “Find a Doctor” tool provided by the American Association for Pediatric Ophthalmology and Strabismus (AAPOS)^43^, as validated by other prior studies.^29^ In case of disagreement, the number provided by the AAO search tool was used. This data, representing the number of pediatric ophthalmologists (“Pediatric Ophthalmology & Strabismus”) per state, was normalized against the population figures from the 2021 State Census Bureau to calculate the physician density per 100,000 inhabitants.^44^ The Relative Demand Index (RDI) was then calculated by dividing the RSV by the pediatric ophthalmologist density, with a separate RDI classification assigned to states devoid of any pediatric ophthalmologists indicating this status.^45^

Incorporated into the study were additional datasets from the US Census Bureau and the Center for Disease Control (CDC) to assess poverty levels, urbanization, age, vision screening rates, and strabismus prevalence state by state.^46–50^ Poverty metrics were evaluated based on the proportion of individuals residing below 50% of the national poverty level.^48^ Each state’s urban or rural designation, alongside age-related demographics, were cataloged, facilitating a nuanced examination of the population structure.^47,49^ Strabismus prevalence data, along with vision screening frequencies, were derived from insurance claims, as reported by the CDC via the Vision and Eye Health Surveillance System (VEHSS)^50^, providing a direct insight into the epidemiological aspects of eye health.^51^

These multifaceted data sources enabled a comprehensive evaluation of the intersections between socioeconomic backgrounds, healthcare accessibility, demographic factors, and eye health outcomes at the state level. By integrating search trends with pediatric ophthalmology service distribution, alongside sociodemographic and health surveillance data, this study offers an expansive overview of the determinants influencing eye health care demand and provision within the United States.

### Statistical Analysis

Using these state-level data, the relationships between search volumes, specialist availability, vision screening, socioeconomics, and demographic factors were explored. The correlation between the relative search volume for pediatric ophthalmology and the availability of pediatric ophthalmologists was first evaluated to demonstrate public interest in pediatric eye health issues relative to the accessibility of specialized care providers. To quantify the nature and extent of these relationships, correlation coefficients (R and R^2^) were calculated to assess the strength and direction of linear associations. Corresponding p-values were also determined to judge statistical significance.

Correlations between the density of pediatric ophthalmologists (adjusted for state population) and the previously described socioeconomic and demographic factors were also evaluated. These factors included state poverty estimates, captured as the proportion of the state population living at or below 50 percent of the poverty threshold, urbanization, prevalence of strabismus, and rates of vision screening. This analysis evaluated how the distribution of pediatric eye care professionals correlates with factors such as economic hardship, urban versus rural residency, and key pediatric eye health indicators. Correlations for additional metrics, including ‘Relative Search Volume’ and ‘Relative Demand Index’, were each assessed against the same socioeconomic and health variables mentioned above.

## RESULTS

### Summary data

Data from all sources were compiled into a single reference table (Table 1). There was significant variability in relative search volumes for pediatric ophthalmology across different states, reflecting diverse levels of public interest and concern regarding pediatric eye health issues (Figure 1). Alaska demonstrated the highest public interest as measured by the relative search volume for strabismus, whereas Arizona showed a more modest RSV of 33, suggesting lower interest in pediatric eye health information.

**Table 1:** All Google trend, pediatric ophthalmologist density, and other demographic and socioeconomic data, displayed per state.

| State | Relative Search Volume | POaS Specialists | Specialist Density | Normalized RDI | Population Under Age 5 | Rate of Vision Screening | Strabismus Prevalence (%) | Urbanization Index | Poverty Index |
| --- | --- | --- | --- | --- | --- | --- | --- | --- | --- |
| Alabama | 40 | 7 | 0.137 | 0.687 | 0.056 | 0.050 | 1.580 | 0.673 | 0.073 |
| Alaska | 100 | 3 | 0.409 | 0.575 | 0.063 | 0.070 | 2.320 | 0.668 | 0.053 |
| Arizona | 33 | 13 | 0.175 | 0.444 | 0.053 | 0.460 | 1.680 | 0.721 | 0.063 |
| Arkansas | 38 | 4 | 0.130 | 0.686 | 0.058 | 0.020 | 1.200 | 0.664 | 0.080 |
| California | 39 | 87 | 0.223 | 0.411 | 0.054 | 0.310 | 1.330 | 0.680 | 0.059 |
| Colorado | 31 | 22 | 0.374 | 0.195 | 0.052 | 0.180 | 1.580 | 0.689 | 0.047 |
| Connecticut | 45 | 12 | 0.332 | 0.319 | 0.049 | 0.330 | 3.480 | 0.773 | 0.049 |
| Delaware | 36 | 4 | 0.388 | 0.219 | 0.053 | 0.270 | 2.090 | 0.722 | 0.046 |
| Florida | 36 | 50 | 0.221 | 0.383 | 0.050 | 0.100 | 1.580 | 0.760 | 0.059 |
| Georgia | 29 | 17 | 0.154 | 0.443 | 0.057 | 0.130 | 1.570 | 0.645 | 0.061 |
| Hawaii | 39 | 8 | 0.557 | 0.165 | 0.055 | 0.140 | 2.320 | 0.686 | 0.053 |
| Idaho | 36 | 2 | 0.102 | 0.832 | 0.058 | 0.040 | 1.150 | 0.568 | 0.045 |
| Illinois | 37 | 45 | 0.359 | 0.243 | 0.054 | 0.050 | 2.140 | 0.793 | 0.061 |
| Indiana | 35 | 9 | 0.131 | 0.628 | 0.058 | 0.120 | 1.260 | 0.706 | 0.061 |
| Iowa | 41 | 7 | 0.218 | 0.442 | 0.056 | 0.030 | 2.020 | 0.681 | 0.052 |
| Kansas | 37 | 4 | 0.136 | 0.640 | 0.060 | 0.190 | 1.700 | 0.726 | 0.055 |
| Kentucky | 39 | 7 | 0.155 | 0.594 | 0.058 | 0.100 | 1.700 | 0.673 | 0.073 |
| Louisiana | 37 | 12 | 0.262 | 0.332 | 0.059 | 0.060 | 1.900 | 0.743 | 0.083 |
| Maine | 45 | 2 | 0.143 | 0.739 | 0.044 | 0.010 | 1.770 | 0.697 | 0.048 |
| Maryland | 39 | 32 | 0.518 | 0.177 | 0.057 | 0.410 | 2.970 | 0.720 | 0.048 |
| Massachusetts | 41 | 31 | 0.443 | 0.218 | 0.049 | 0.270 | 2.610 | 0.768 | 0.051 |
| Michigan | 35 | 22 | 0.219 | 0.376 | 0.053 | 0.140 | 2.430 | 0.776 | 0.064 |
| Minnesota | 39 | 18 | 0.314 | 0.293 | 0.057 | 0.160 | 1.900 | 0.695 | 0.044 |
| Mississippi | 37 | 4 | 0.136 | 0.640 | 0.058 | 0.040 | 0.830 | 0.647 | 0.085 |
| Missouri | 41 | 14 | 0.226 | 0.427 | 0.057 | 0.090 | 1.470 | 0.742 | 0.061 |
| Montana | 37 | 4 | 0.353 | 0.247 | 0.051 | 0.120 | 1.240 | 0.649 | 0.056 |
| Nebraska | 43 | 5 | 0.253 | 0.401 | 0.062 | 0.090 | 1.970 | 0.688 | 0.050 |
| Nevada | 34 | 6 | 0.188 | 0.426 | 0.054 | 0.110 | 0.830 | 0.711 | 0.064 |
| New Hampshire | 45 | 6 | 0.428 | 0.248 | 0.045 | 0.110 | 2.330 | 0.684 | 0.034 |
| New Jersey | 35 | 33 | 0.355 | 0.232 | 0.055 | 0.350 | 2.820 | 0.746 | 0.044 |
| New Mexico | 38 | 6 | 0.284 | 0.315 | 0.050 | 0.070 | 1.560 | 0.747 | 0.077 |
| New York | 41 | 79 | 0.404 | 0.239 | 0.054 | 0.340 | 3.970 | 0.769 | 0.073 |
| North Carolina | 36 | 23 | 0.212 | 0.399 | 0.055 | 0.350 | 1.510 | 0.657 | 0.057 |
| North Dakota | 50 | 0 | 0.000 | Max | 0.061 | 0.040 | 0.950 | 0.637 | 0.054 |
| Ohio | 40 | 34 | 0.288 | 0.326 | 0.056 | 0.060 | 2.680 | 0.781 | 0.063 |
| Oklahoma | 35 | 11 | 0.271 | 0.304 | 0.060 | 0.020 | 1.610 | 0.680 | 0.071 |
| Oregon | 32 | 11 | 0.260 | 0.290 | 0.047 | 0.260 | 1.710 | 0.710 | 0.058 |
| Pennsylvania | 48 | 40 | 0.309 | 0.366 | 0.052 | 0.220 | 2.430 | 0.763 | 0.055 |
| Rhode Island | 44 | 5 | 0.456 | 0.227 | 0.048 | 0.170 | 1.940 | 0.805 | 0.050 |
| South Carolina | 36 | 10 | 0.186 | 0.455 | 0.053 | 0.060 | 1.920 | 0.663 | 0.064 |
| South Dakota | 41 | 3 | 0.326 | 0.296 | 0.063 | 0.070 | 1.290 | 0.614 | 0.055 |
| Tennessee | 37 | 12 | 0.168 | 0.517 | 0.057 | 0.240 | 1.670 | 0.654 | 0.060 |
| Texas | 39 | 42 | 0.138 | 0.667 | 0.063 | 0.520 | 1.430 | 0.599 | 0.064 |
| Utah | 38 | 6 | 0.176 | 0.510 | 0.068 | 0.150 | 1.270 | 0.535 | 0.038 |
| Vermont | 44 | 0 | 0.000 | Max | 0.043 | 0.120 | 1.750 | 0.663 | 0.042 |
| Virginia | 37 | 29 | 0.333 | 0.262 | 0.055 | 0.130 | 1.920 | 0.668 | 0.052 |
| Washington | 33 | 15 | 0.192 | 0.405 | 0.054 | 0.090 | 1.890 | 0.674 | 0.053 |
| West Virginia | 48 | 2 | 0.113 | 1.000 | 0.049 | 0.070 | 1.700 | 0.738 | 0.080 |
| Wisconsin | 34 | 15 | 0.254 | 0.315 | 0.052 | 0.060 | 1.710 | 0.739 | 0.049 |
| Wyoming | 26 | 0 | 0.000 | Max | 0.052 | 0.020 | 0.940 | 0.727 | 0.048 |

POaS - Pediatric Ophthalmology and Strabismus.
RDI - Relative Demand Index

**Figure 1.**
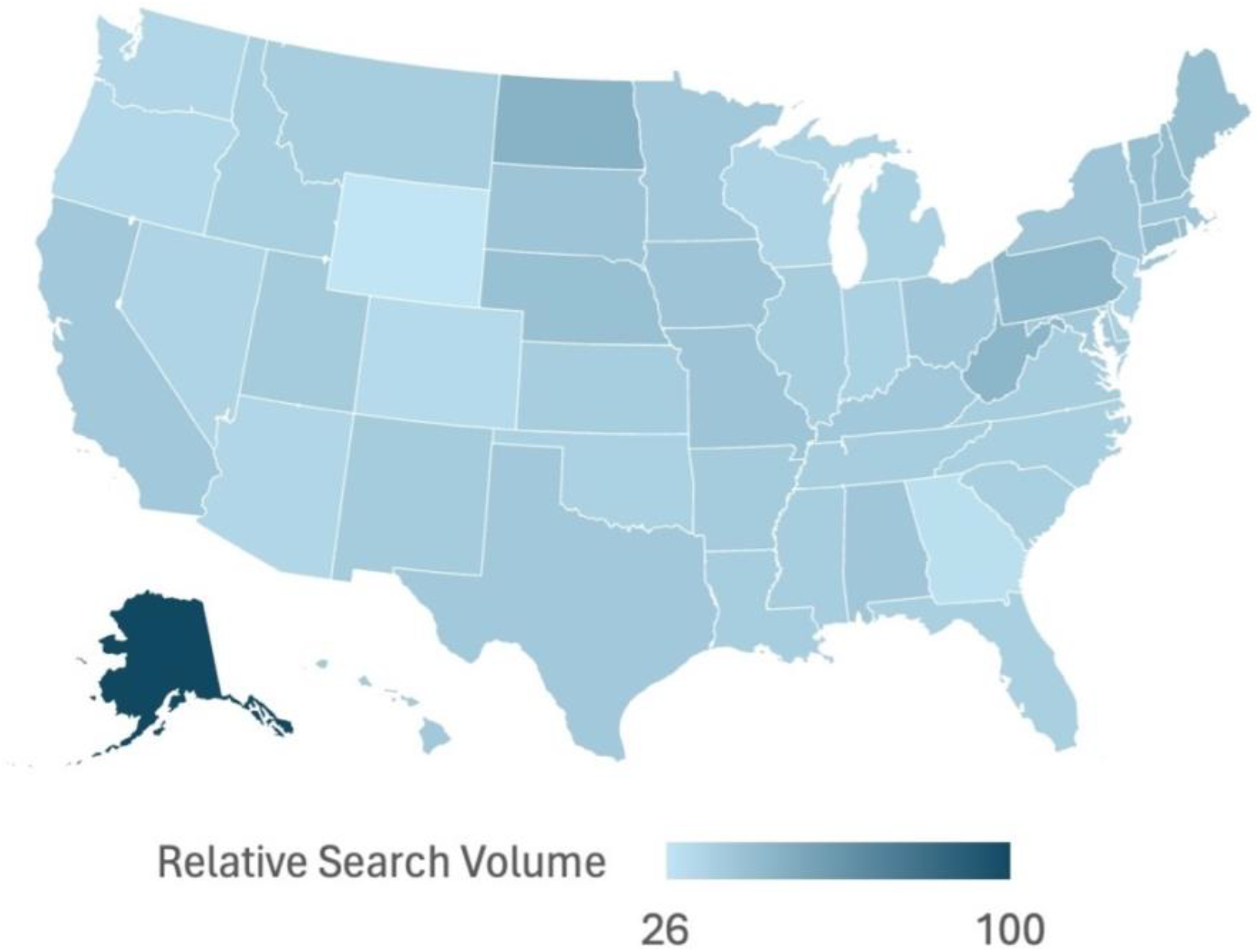
Google Trends Relative Search Volumes (RSVs) for search term “strabismus” from 1/1/14 - 1/31/23 displayed for each state, with increased color density corresponding to increased RSV.

Analysis of the American Academy of Ophthalmology (AAO) search tool data highlighted notable disparities in the number of pediatric ophthalmology and strabismus specialists available across states. This variation underscores the uneven distribution of specialized pediatric eye care professionals, potentially impacting the accessibility and quality of care for children’s eye health. For example, California, with 87 specialists, stands out for its high specialist density, contrasting with states like Alaska, which has only 3 specialists, despite a high public interest in pediatric eye health.

Calculating normalized RDI from RSV and specialist density reveals notable differences across various states (Figure 2). In addition to North Dakota, Vermont, and Wyoming, which have no pediatric ophthalmologists, West Virginia stands out with the highest normalized RDI value of 1.0, indicating high demand relative to its population size. In contrast, Hawaii registers the lowest with a value of approximately 0.165, showing the least demand in comparison to other states. On average, the states have a normalized RDI of about 0.416, with the median at roughly 0.383.

**Figure 2.**
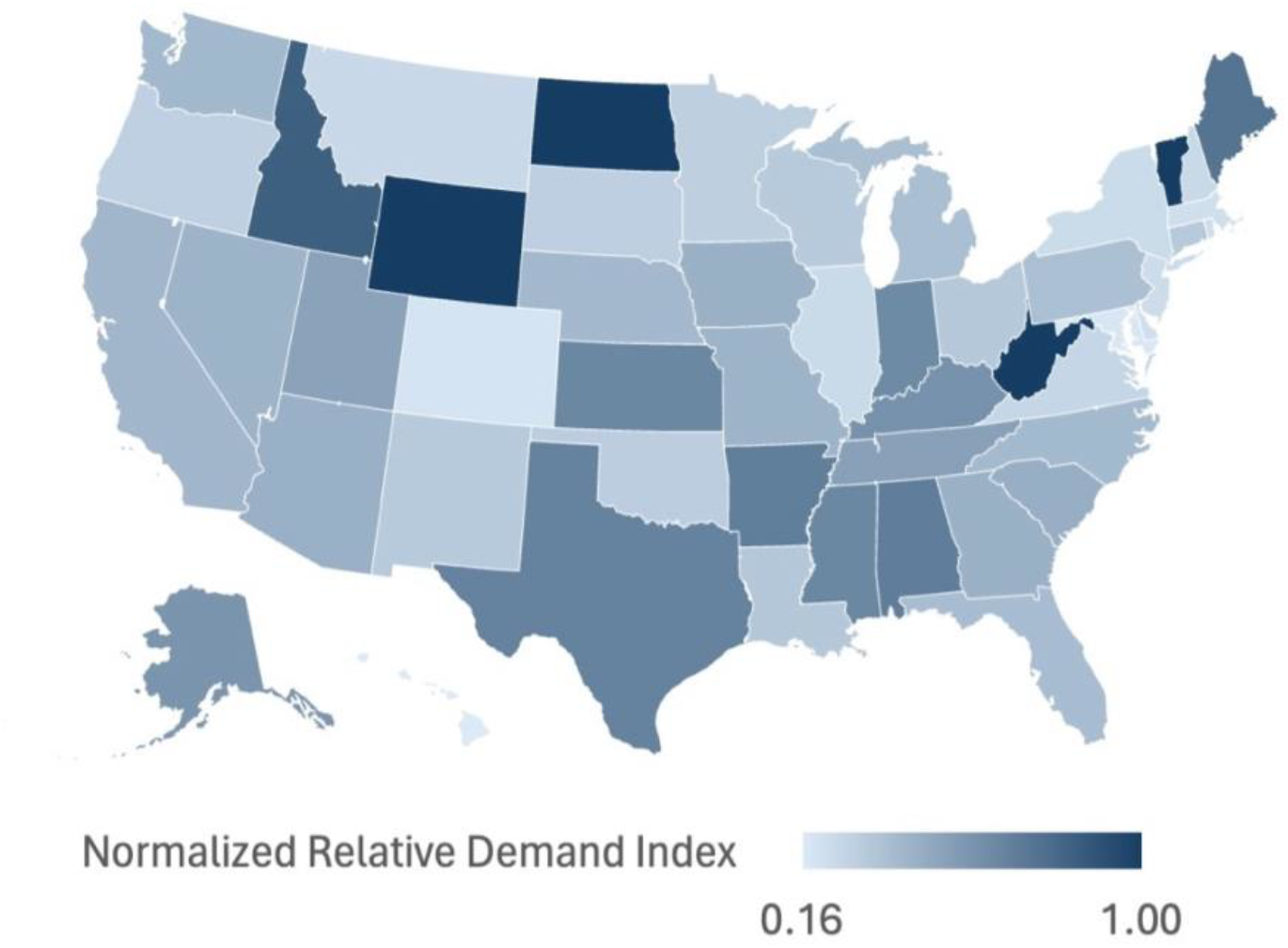
Normalized Relative Demand Index (RDI) as calculated from Relative Search Volume (RSV) and specialist density, with increased color density corresponding to increased RDI.

The Census Bureau data provided critical insights into urbanization, poverty, and the proportion of the population under age 5 across states. These socioeconomic and demographic variables play a significant role in shaping the demand for pediatric ophthalmology services. The data reveals a complex landscape of urban versus rural residency, varying poverty levels, and the proportion of the population under age 5, all of which are crucial factors influencing access to and the need for pediatric eye care services.

Lastly, data from the VEHSS on strabismus prevalence and vision screening rates offered valuable perspectives on the state of pediatric eye health across the country (Figure 3). The prevalence of strabismus, a common condition affecting eye alignment in children, varied across states, as did the rates of vision screening, an essential preventive measure to detect eye health issues early. These metrics, when considered alongside Google trends data and specialist availability, highlight the intricate interplay between public health efforts, disease prevalence, and healthcare resource allocation in addressing pediatric eye health needs.

**Figure 3.**
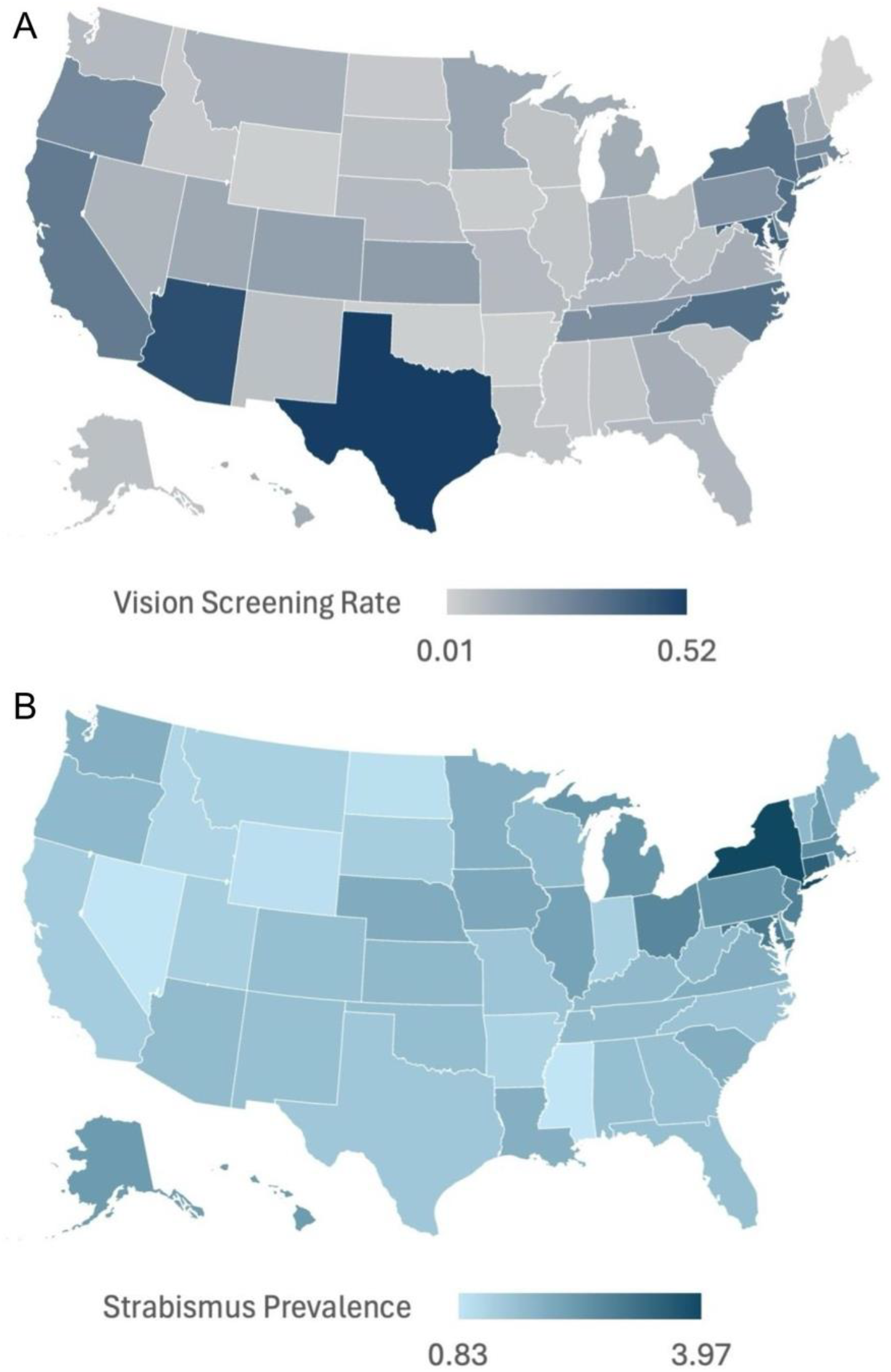
Insurance claims data as reported by the CDC displayed per state, with increased color density corresponding to increased rate or prevalence for **A)** Patient screening rates, captured as percent of total population each year and **B)** Strabismus prevalence (%) per state, for population under age 18.

### Associations with searches for strabismus

In the analysis of state-level data regarding ophthalmology and public health metrics, several key findings emerged (Table 2).

**Table 2:**
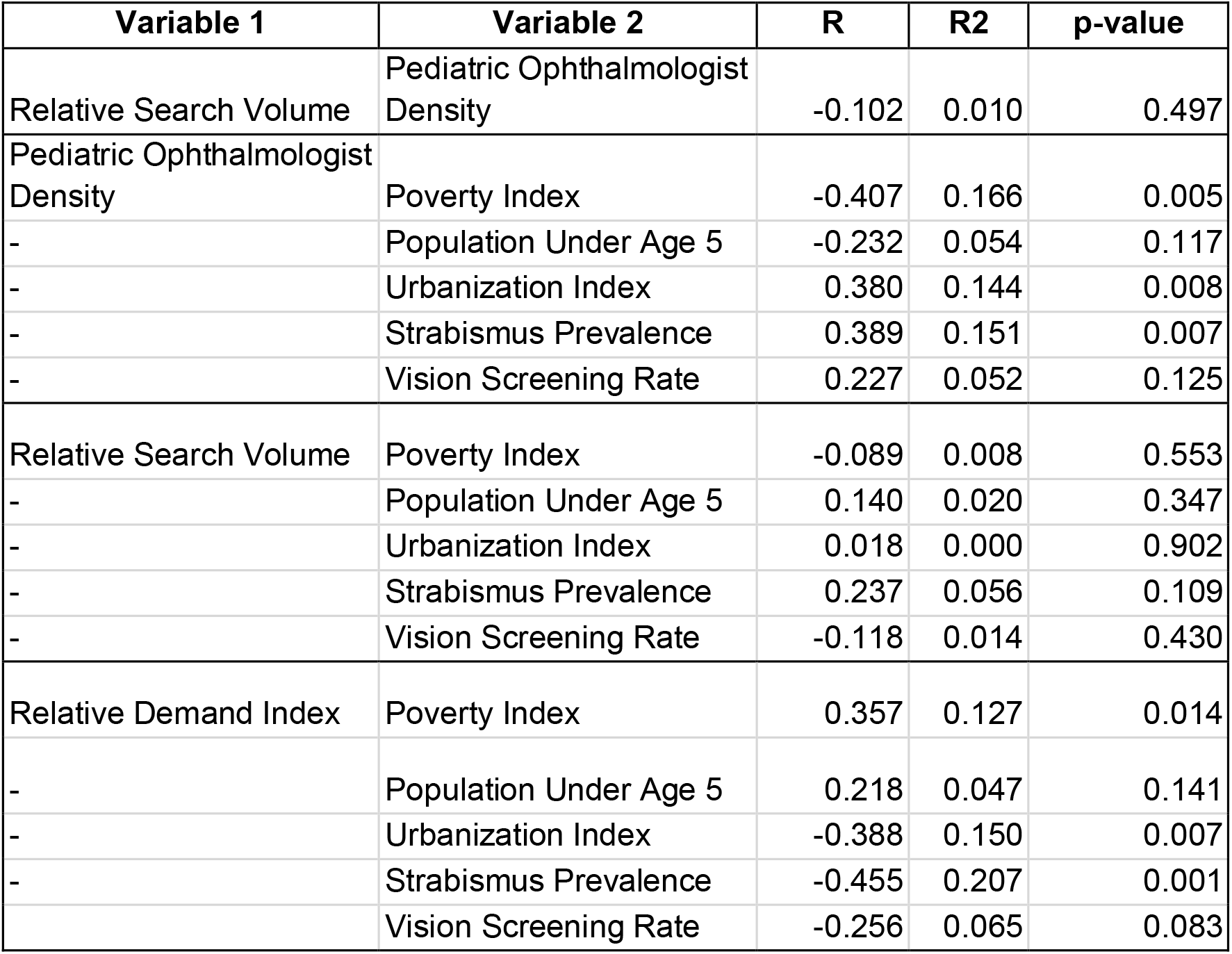
Associations with Pediatric Ophthalmologist Density, Relative Search Volume, and Relative Demand Index. All calculated correlation coefficients (R), determination coefficients (R2), and associated p-values.

| Variable 1 | Variable 2 | R | R2 | p-value |
| --- | --- | --- | --- | --- |
| Relative Search Volume | Pediatric Ophthalmologist Density | -0.102 | 0.010 | 0.497 |
| Pediatric Ophthalmologist Density | Poverty Index | -0.407 | 0.166 | 0.005 |
| - | Population Under Age 5 | -0.232 | 0.054 | 0.117 |
| - | Urbanization Index | 0.380 | 0.144 | 0.008 |
| - | Strabismus Prevalence | 0.389 | 0.151 | 0.007 |
| - | Vision Screening Rate | 0.227 | 0.052 | 0.125 |
| Relative Search Volume | Poverty Index | -0.089 | 0.008 | 0.553 |
| - | Population Under Age 5 | 0.140 | 0.020 | 0.347 |
| - | Urbanization Index | 0.018 | 0.000 | 0.902 |
| - | Strabismus Prevalence | 0.237 | 0.056 | 0.109 |
| - | Vision Screening Rate | -0.118 | 0.014 | 0.430 |
| Relative Demand Index | Poverty Index | 0.357 | 0.127 | 0.014 |
| - | Population Under Age 5 | 0.218 | 0.047 | 0.141 |
| - | Urbanization Index | -0.388 | 0.150 | 0.007 |
| - | Strabismus Prevalence | -0.455 | 0.207 | 0.001 |
|  | Vision Screening Rate | -0.256 | 0.065 | 0.083 |

A moderate positive correlation was observed between the number of ophthalmologists and urbanization (R = 0.302, p = 0.033), indicating that more ophthalmologists are available in states with higher levels of urbanization. Furthermore, the analysis revealed a strong positive correlation between the number of ophthalmologists and the vision screening rate (R = 0.486, p = 0.0003), suggesting that the availability of ophthalmologists within states is associated with higher rates of vision screening.

The study also found a moderate negative correlation between the relative demand index and urbanization (R = −0.388, p = 0.007), displaying how the demand for ophthalmological services per pediatric ophthalmologist inversely relates to urbanization levels. Moreover, a moderate negative correlation between the relative demand index and the strabismus prevalence (R = −0.455, p = 0.001) was observed, indicating that higher demand for ophthalmological services per ophthalmologist is associated with lower prevalence of strabismus.

Another notable finding from this analysis is the weak and statistically non-significant correlation between the relative search volume for ophthalmological services and the poverty estimate, defined as 50 percent of the poverty level (R = −0.081, p = 0.577). This suggests that the search volume for these services does not strongly correlate with the poverty level across states.

## DISCUSSION

Overall, this analysis underscores the complex relationship between public interest in pediatric eye health, the distribution of healthcare resources, socioeconomic factors, and health outcomes. By evaluating a total of 16 distinct pairings, this comprehensive approach provided nuanced insights into the dynamics between public demand for information on strabismus, the supply of healthcare resources, and various demographic and health factors. The methodology of this study offered a structured framework to assess the interconnections among healthcare resource distribution, public interest, and demographic and health variables in pediatric ophthalmology.

The observed variance in search volumes suggests differences in awareness or perceived importance of pediatric eye health across regions. The relationship between urbanization and pediatric ophthalmologist availability relationship also underscores the role of urbanization in access to health care. The negative correlation between relative demand index and urbanization further indicates that urban areas, with their higher concentration of healthcare resources, experience relatively lower demand per provider compared to less urbanized areas. The positive association between availability of ophthalmologists within states and higher rates of vision screening emphasizes the importance of healthcare accessibility. The finding of a negative correlation between the relative demand index and the strabismus prevalence could similarly reflect both underdiagnosis due to a shortage of pediatric ophthalmologists and the impact of healthcare accessibility and resource distribution on managing and reducing the incidence and therefore prevalence of specific health conditions.

These findings provide insights into the distribution and demand for ophthalmological services across different states, highlighting the influence of urbanization and the availability of healthcare professionals on public health outcomes. Demographic factors, varying state by state, play a crucial role in the management and outcomes of strabismus, influencing access to care, treatment adherence, and the ability to afford necessary interventions. Addressing these disparities requires a multifaceted approach, including policy changes, increased funding for public health programs, and community-based initiatives aimed at reducing barriers to eye care. There remains a need for targeted public health interventions and resource allocation strategies to bridge gaps in care and improve pediatric eye health outcomes across diverse communities. Further research is warranted to continue to explore these relationships and their implications for healthcare policy and resource allocation.

Limitations of this study include Google searches for “strabismus” not for the purpose of seeking care; a portion of search volumes may originate from students, researchers, healthcare providers, or individuals with other interest in strabismus. Additionally, the AAO “Find an Ophthalmologist” tool relies on ophthalmologists to self-report their field of practice and location and may be incomplete. Though we use “strabismus” Google relative search volume as a proxy for interest and demand for pediatric ophthalmologists, strabismus is not limited to the pediatric population. Moreover, a decade worth of Google search volumes was analyzed in this study, while trends and demands may fluctuate year-to-year and therefore the relative search volumes spanning the decade may not reflect the relative search volumes in the most recent years. It should also be noted that Google trends search volume data is dynamically updated and subject to change as Google maintains this database. Also, operationalizing demand through search volume assumes that the need or desire for ophthalmologic services is accurately reflected by relative Internet search volume, which may not hold true across different populations. For example, the decrease in adjusted search rates in non-urban areas may not necessarily indicate reduced demand, but could be attributed to different healthcare-seeking behaviors among non-urban populations, among other potential latent structural differences.

Technological advancements present new opportunities for improving strabismus care. Recent developments in teleophthalmology, digital screening tools, and surgical innovations offer promising avenues for enhancing access to diagnostic and treatment services. ^52–54^ Exploring the integration of these technologies into current care models could address some of the challenges related to healthcare access and the timely management of strabismus.

Strabismus represents a significant public health issue that requires timely and effective intervention to prevent long-term visual impairment. The challenges faced by pediatric ophthalmology, including issues related to underpayment and the resulting unfilled need for specialized care, compound the difficulties in managing this condition. This research addresses a gap in the scientific literature regarding the demand for pediatric ophthalmologists’ services such as strabismus correction as captured by online search volume, and how this relates to physician availability, alongside various demographic and health factors, on a state-by-state basis.

## Data Availability

All data produced are available upon reasonable request to the authors

https://data.census.gov/table/ACSST1Y2022.S0101?q=population

https://data.census.gov/table/ACSST1Y2022.S1701?q=poverty

https://data.census.gov/table/DECENNIALCD1162010.H2q%3Durbanization&sa=D&source=docs&ust=1711327368394090&usg=AOvVaw2256qJPf0TtFb603U516DH

https://www.cdc.gov/visionhealth/vehss/project/index.html

https://secure.aapos.org/i4a/memberDirectory/index.cfm?directory_id=7&pageID=3322

https://secure.aao.org/aao/find-ophthalmologist

## Acknowledgement

This work is supported by National Institutes of Health/National Eye Institute Grants (R01EY034148, R01EY029058, R01EY11008, R01EY19869, R01EY027510, R01EY026574, EY018926, P30EY022589, K99EY030942, R01EY034146, T35EY033704, DP5OD029610); and an unrestricted grant from Research to Prevent Blindness (New York, NY). The sponsor or funding organization had no role in the design or conduct of this research.

